# Vaccine Hesitancy, Coercion and Regret: Post-Pandemic Lessons on COVID-19 Vaccine Policies and Outreach among Newcomer Communities in Alberta, Canada

**DOI:** 10.1101/2025.02.03.25320914

**Authors:** Minnella Antonio, Edna Ramirez-Cerino, Adanech Sahilie, Mussie Yemane, Michael Youssef, Ingrid Nielssen, Linda Holdbrook, Mohammad Yasir Essar, Maria Santana, Denise Spitzer, Kevin Pottie, Gabriel E. Fabreau

**Affiliations:** Refugee Health YYC O’Brien Institute for Public Health, Cumming School of Medicine, University of Calgary, Calgary, Alberta, Canada; University of Alberta, School of Public Health, Edmonton, Alberta, Canada; Department of Community Health Sciences, Cumming School of Medicine, Calgary, Alberta, Canada; Alberta Strategy for Patient-Oriented Research (SPOR) SUPPORT Unit (AbSPORU), Patient Engagement Team, Calgary, Alberta, Canada; Patient and Community Engagement Research Program, University of Calgary, Continuing Education, Calgary, Alberta, Canada; Departments of Pediatrics and Community Health Sciences, Cumming School of Medicine, University of Calgary, Calgary, Alberta; Department of Family Medicine, Cumming School of Medicine, University of Calgary, Calgary, Alberta, Canada; Department of Medicine, Cumming School of Medicine, University of Calgary, Calgary, Alberta, Canada; Western University Department of Family Medicine, Dalhousie University

**Keywords:** COVID-19, vaccines, newcomers, migrant health, qualitative study, patient-oriented research, Community based participatory research, public health communication

## Abstract

**Background:** Since the COVID-19 pandemic routine vaccination rates have dropped in Canada. Many newcomers and refugees experience significant vaccine inequities despite wide vaccine availability and COVID-19 pandemic vaccination campaigns. We aimed to investigate post-pandemic vaccine hesitancy, acceptance, and vaccine outreach strategies among newcomers’ communities.

**Methods:** We conducted a prospective community-based-participatory research (CBPR) qualitative study with self-identified newcomers in Alberta between October 2022 - January 2023. Community Scholars, leaders representing various ethnocultural communities trained in community-engaged research conducted semi-structured interviews and focus groups in English and first languages. Scholars collected and translated detailed qualitative notes and sociodemographic data with a standardized survey. Qualitative data was thematically analyzed and coded using a consensus-based approach.

**Results:** We conducted two focus groups and five semi-structured interviews with 12 participants, 50% who identified as female and originated from the Philippines, Ethiopia, Eritrea, Mexico, Burundi, and Egypt. We identified three main themes, each with two subthemes: (1) vaccine hesitancy due to lack of reliable information and religious and cultural beliefs, (2) access to COVID-19 vaccines and information, and (3) vaccine acceptance as voluntary or coerced. Employer mandated vaccination emerged as a critical issue with potential long-term negative public health implications, leading to vaccine regret and loss of trust of public health authorities and healthcare systems.

**Conclusion:** During population-wide COVID-19 vaccination campaigns newcomers perceived their communities’ circumstances were overlooked, potentially increasing vaccine hesitancy. Perceived coercive vaccination policies had unintended negative public health consequences. These findings may help inform future emergency and routine public health vaccination policies.

## Introduction

> *“Sometimes immigrants feel like okay, probably the health services, the family doctors, the pharmacists, the vaccines are only for Canadians. So, we have to change that …, they said to me … ‘I’m not going to ask for that kind of things because are only for Canadians. Are not for immigrants. Are not for Mexicans. Are not for us.” (Interviewee A)*

The COVID-19 pandemic exacerbated longstanding health inequities, particularly among newcomer populations compared to the general Canadian population (Crawshaw et al., 2021). Despite rapidly implemented population-based COVID-19 vaccination programs in Canada, migrants and refugees experienced disproportionately higher rates of infection, morbidity, and mortality compared to the general population (Crawshaw et al., 2021). These disparities are likely attributable to systemic barriers to healthcare access, including language barriers, limited access to reliable health information, and greater vulnerability to mis and disinformation (Kalich et al., 2016). Collectively, these barriers contributed to lower vaccination rates among newcomer communities compared to non-immigrant populations in Canada (MacDonald et al., 2022). Responding to these challenges, community-based participatory research (CBPR) and community-engaged outreach strategies have shown promise to improve vaccine uptake among migrant and refugee populations (Holdbrook et al., 2023) (Marquez et al., 2021). However, in Canada, such strategies were deployed piecemeal across jurisdictions during and after the pandemic, leaving knowledge gaps regarding newcomers’ experiences in accessing COVID-19 vaccines and related information. Understanding these experiences is essential to develop equitable public health interventions, especially given Canada’s post-pandemic record immigration levels (Statistics Canada, 2022), and serious decline in routine vaccination rates resulting in multiple outbreaks of vaccine-preventable diseases like measles and pertussis (Vaccines in Canada, 2024).

This study aimed to employ CBPR methods to investigate post-pandemic vaccine hesitancy, acceptance, and vaccine outreach strategies among urban and rural newcomer communities in Alberta. As part of this broader study, we recruited and trained Community Scholars - community leaders from diverse ethnic backgrounds, languages, and cultural competencies, through a formal one year research program (the Patient and Community Engagement Research (PaCER) program) designed to engage community members with lived experience in health research to inform planning and policy around patients’ health conditions (Patient and Community Engagement Research (PaCER) | University of Calgary, n.d.). Community Scholars (CSs) were highly motivated to understand and share the information, needs, and experiences of newcomers accessing COVID-19 vaccine information and vaccines in Alberta during the pandemic. By grounding our inquiry in the perspectives of those most affected, we sought to inform public health strategies that would enhance vaccine equity and health outcomes for newcomer populations.

This study utilized a post-pandemic, post-mortem lens to maximize the learnings from the COVID19 pandemic to inform context-specific and universal recommendations for vaccination and public health outreach strategies among newcomer communities. In doing so, this research contributes to critical public health discourse, emphasizing the need for culturally responsive and community-driven approaches to vaccine outreach and health equity.

## Methods

We conducted a prospective, community-engaged qualitative study using CBPR principles and the PaCER framework (University of Calgary Press, n.d.), designed to prioritize the voices and experiences of marginalized newcomer communities in Alberta, Canada between May 2022 and January 2023. CBPR, rooted in critical social theory, positions participants not as subjects of research but as co-creators of knowledge (Collins et al., 2018). This approach challenges traditional research paradigms by recognizing the expertise of lived experience and promoting collaborative, action-oriented outcomes (Collins et al., 2018). The study team consisted of five Community Scholars (CSs) who collectively spoke Tigrinya, Amharic, Arabic, Spanish, Tagalog, and English. They were embedded within the University of Calgary’s Refugee Health YYC research program and co-sponsored by the Alberta International Medical Graduates Association and led the study from inception to completion (AIMGA, n.d.) (Refugee Health YYC, n.d.). To capture diverse perspectives, we ensured study participants could participate in their first languages throughout all research stages.

### The PaCER Framework

We utilized the PaCER framework, which structures research into three phases—SET, COLLECT, and REFLECT (Figure 1)— to engage community members throughout all study stages. This approach ensures methodological rigour while grounding the research in the lived experiences of newcomer communities (University of Calgary Press, n.d.). In the SET phase, community members with relevant lived experience provided input on the research question, confirming its relevance and helping design the study’s scope. During the COLLECT phase, data was gathered from participants through focus groups and semi-structured interviews, with thematic analysis conducted on detailed notes. Finally, in the REFLECT phase, participants reviewed and validated the emergent themes, offered insights on future research priorities, and contributed to the knowledge translation strategy for their communities (University of Calgary Press, n.d.).

**Figure 1:**
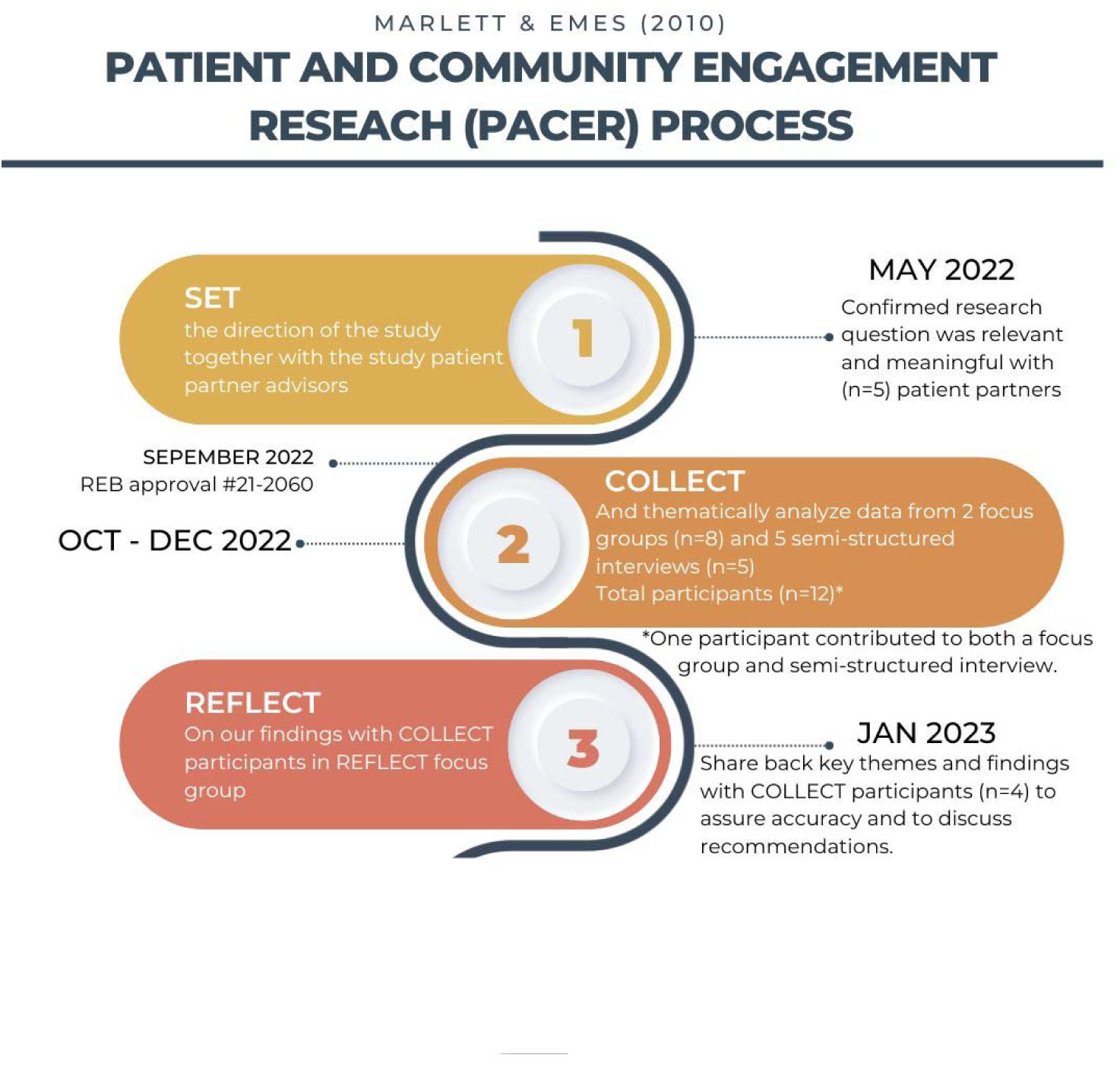
PaCER Process, including study timeline, focus groups, semi-structured interviews, analysis and member checking steps

### Participant Selection and Recruitment

Study eligibility criteria included adults (18 years or older), who were self-identified as newcomers or racialized, lived in Alberta, and interested in participating in a community-led study. Various recruitment methods were employed (eFigure 1), including social media posts, snowball recruitment, and outreach to community organizations; however, these initial efforts were largely unsuccessful. Given the Community Scholars’ (CSs) lived experience as newcomers and their active involvement in newcomer community organizations, they leveraged their strong ties within these networks for study recruitment. To facilitate clear communication, one team member was designated as the primary liaison for participants comfortable speaking English. For those preferring another language, we provided contact information for a team member fluent in their first language. Potential study participants were then sent an email with an invitation letter, study information, consent form, and link to an anonymous demographic survey.

### SET Discussion Group and Study Co-Design

CSs first developed the research question, “Understanding newcomers’ COVID-19 vaccine access and experience,” through an iterative process of literature review and internal discussions. Once defined, CSs recruited community partners for a SET discussion group via Zoom. The SET discussion group and CSs co-designed the research question, exploratory topics, interview and focus group guides to ensure relevance to newcomer communities. The SET group focused on the importance of COVID-19 practices and generated ideas for participant recruitment and interview questions, co-designing the COLLECT phase. Five community members participated, expressing interest in exploring the intersection of vaccine access and information inequity with racism, discrimination, and the mental health impacts of vaccination. The participants also stressed the importance of including socioeconomic and religious diversity in the study population and respecting each participant’s belief systems, acknowledging the sensitivity of race, discrimination, and COVID-19 vaccines.

Using the SET discussion group’s findings, CSs modified the research protocol, focus group and interview guides for the COLLECT phase. The research question was refined to “What are the experiences of newcomers accessing COVID-19 vaccine information and vaccines in Alberta during the COVID-19 pandemic?”

### Data Collection COLLECT Focus Groups and Semi-Structured Interviews

We held two COLLECT focus groups and five semi-structured interviews via the secure University of Calgary Zoom platform (eTable 1). To ensure equitable participation, real-time interpretation was provided in the six languages spoken by CSs. Participants provided written or recorded oral consent and all sessions were audio recorded for note taking purposes only. These detailed notes served as the qualitative data for analysis.

The first focus group was conducted primarily in Amharic, with participants transitioning from English to Amharic as the discussion progressed. were translated into English for analysis. The second focus group included participants from various ethnic backgrounds, all comfortable speaking English, though some chose to speak in their first language, with CS team members providing real-time translations. Individual semi-structured interviews were offered to participants unable to attend focus groups or who preferred to share their experiences privately. Except for one, all interviews were conducted by pairs of interviewers, with at least one interviewer fluent in the participant’s first language, and one note taker. Most participants kept their video on during the interviews.

### Data Analysis

We used an iterative six-step thematic analysis approach developed by Braun and Clarke (2021) to analyze focus group and interview data (Braun & Clarke, 2023). This approach was selected for its flexibility and support for researcher reflexivity. We used Microsoft Excel (Seattle, WT) to sort and analyze the data, using a separate sheet for the notes from each interview or focus group and a single spreadsheet cell for each collective thought or idea expressed by participants. The CS team iteratively developed a codebook, starting with codes from the first interview and adding new codes as additional data were analyzed, applying this codebook to the remaining focus group and interview data adding new codes as they emerged.

Each focus group and interview were double coded. A master codebook was created, and pivot tables were used to aggregate the most frequently used codes (eTable 2). Focus group and interview data were summarized and presented to participants in a REFLECT focus group as part of a member-checking process. This step allowed participants to clarify or refine emerging themes (University of Calgary Press, n.d.).

## REFLECT Focus Group

Following the identification of key themes, subthemes, and recommendations, COLLECT participants were invited to a final REFLECT focus group via Zoom (Figure 1, eTable 3). Anonymity was reiterated, and participants remained actively engaged via video, the “raise hand” function, or the chat box. The study findings were presented, and participants were asked if the results accurately reflected their experiences. We also asked clarifying questions that arose during the focus groups and interviews such as, “What sources of information do you trust the most?” and, “How do you determine the reliability of information?” To validate our findings, we asked questions such as, “Do you think language barrier, cultural, socioeconomic status and religious background influence vaccine hesitancy?”

### Ethical Considerations

The study was approved by the University of Calgary research ethics board (REB21-2060). The study followed the Consolidated Criteria for Reporting Qualitative (COREQ) Research guidelines (Tong, Sainsbury, & Craig, 2007) (eTable 4) and the Guidance for Reporting Involvement of Patients and the Public (GRIPP) guidelines (Staniszewska et al., 2017) (eTable 5)

## Results

We conducted one discussion group, three focus groups (one SET, two COLLECT, and one REFLECT), and five semi-structured interviews, including one participant who had previously attended a focus group and had further experiences to share (Figure 1). Focus groups ranged from 70-92 minutes and interviews from 35-90 minutes.

### Participant Characteristics

Twelve participants were included in the study, with equal gender representation. Participants self-identified their gender and country of origin during focus groups and interviews (Table 1).

**Table 1:** Focus group and interview participants’ characteristics.

| Participant Country<br>of Origin | Male<br>N=6 (%) | Female<br>N=6 (%) | Total<br>N=12 (%) |
| --- | --- | --- | --- |
| Philippines | 1 (16.7) | 3 (50) | 4 (33) |
| Ethiopia | 3 (50) | 0 (0) | 3 (25) |
| Eritrea | 1 (16.7) | 1 (16.7) | 2 (17) |
| Mexico | 0 (0) | 1 (16.7) | 1 (8.3) |
| Burundi | 1 (16.7) | 0 (0) | 1 (8.3) |
| Egypt | 0 (0) | 1 (16.7) | 1 (8.3) |

### Themes, Subthemes and Exemplar Quotes

The analysis revealed three main themes, and six subthemes related to vaccine hesitancy, accessibility, and acceptance among newcomers (Figure 2). The most frequent codes identified were “trusted sources of information,” “vaccine accessibility,” and “vaccine experience” (eTable 2).

**Figure 2:**
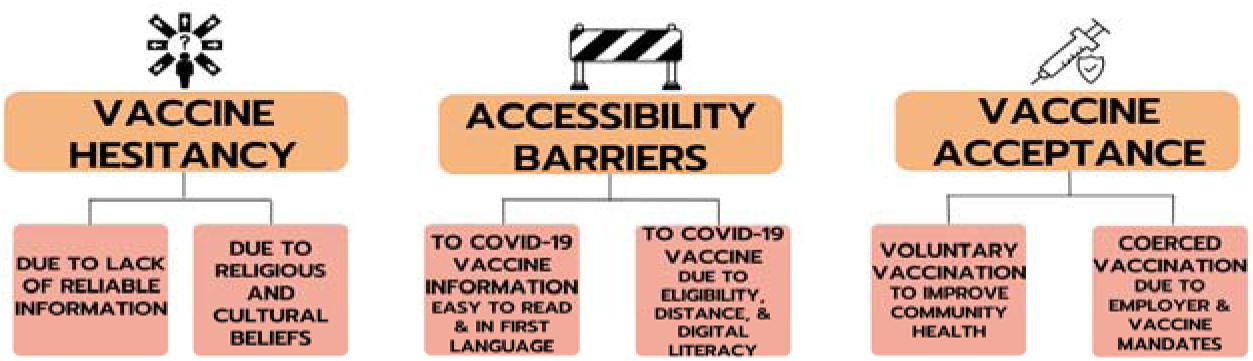
Emergent themes and subthemes regarding vaccine hesitancy, vaccine acceptance and vaccine accessibility for newcomers in Alberta seeking COVID-19 vaccines and COVID-19 vaccine information.

#### 1. Vaccine Hesitancy

Participants described vaccine hesitancy as a major issue among newcomer communities, with two underlying sub-themes: vaccine hesitancy due to lack of reliable information and hesitancy influenced by religious and cultural beliefs.

### Vaccine Hesitancy Due to Lack of Reliable Information

Participants reported facing challenges in processing the overwhelming volume of medical information from various sources, such as global and local news, faith-based leaders, social media, and trusted individuals. This often conflicting and evolving information caused confusion and fear, contributing to COVID-19 vaccine hesitancy.

> *“It was at times overwhelming because it was just a lot of information. If you were on social media there was a lot of misinformation that your brain, just, to constantly like filter out, you know, what’s true and what’s not… , again it was just overwhelming at times, it could be very confusing because of the constantly changing or being updated information.” (Interviewee B)*

Five participants noted that the “infodemic” of COVID-19-related misinformation negatively impacted their or their community’s decision-making about vaccination (Table 2). Early in the pandemic this infodemic, paired with a perceived urgency for those whose employment, and only income source, relied on receiving two COVID-19 vaccine doses, reduced participants’ confidence and increased their uncertainty about the vaccine’s safety. For three participants, employer vaccine mandates were the only reason they chose to be immunized, feeling they lacked sufficient time to make informed decisions (Table 2). When asked if they would have received the vaccine without employer mandates, two participants said no: “*I don’t think so. It’s fifty-fifty position, but it’s just my own belief.” (Interviewee C)*

**Table 2.** COVID-19 vaccine hesitancy and underlying motivators among newcomer communities.

| Theme | Sub-Theme | Quote |
| --- | --- | --- |
| Vaccine Hesitancy | Vaccine Hesitancy Due to Lack of Reliable Information | *"Since it [the COVID-19 vaccine's creation] has been a rush we all know, we didn't know that what could be the effect of this vaccine. So yeah, at first, I was really scared, but I was able to get it because I don't have a choice, I need it for my work." - (Interviewee C)* |
|  |  | *"There's nice doctor in our church also she called, um I think [redacted]? She's also member in [agency]. She did like a meeting for us at the church to give information about* |
|  |  | *the pandemic and the COVID vaccine and what can we do as a member of the community to help all the people around us, you know? So and she did also I think a zoom meeting maybe one time or two times with [redacted] to give all the people interested to get information. So it was so helpful and I think this was like a step for us to go search for the vaccine like okay we trust it now so how we get it? So it was a first step." - (Interviewee I)* |
|  | Vaccine Hesitancy Due to Religious and Cultural Beliefs | *"As we all know, Ethiopian and Eritrean communities are religious people...The impact that religion had brought about a great mess in combating this disease is beyond proportion because it has been widely propagated that the vaccine itself is a tool of controlling the pious religious people by few who tend to dominate society through non-religious ways – through material ways." - (Interviewee K)*<br>*"Because I understand English good, so language wasn't a barrier, as well as religious [beliefs], as I am Coptic Orthodox, and we totally agree with vaccination to protect ourselves. In the Egyptian culture, we agree with the vaccine as well." - (Interviewee I)* |

### Vaccine Hesitancy Due to Religious and Cultural Beliefs

For some newcomer communities, religious and cultural beliefs were significant factors influencing vaccine acceptance, particularly in Filipino, Burundian, Ethiopian, and Eritrean communities. Participants reported mixed religious perspectives, some citing the close intertwining of religion and culture with COVID-19 vaccine denouncements by members of their ethnocultural communities (Table 2). Additionally, some participants experienced feelings of stigma after immunization, as members of their communities questioned and argued against their decisions to get vaccinated.

> *“For myself, I didn’t tell my mother that I want to get the vaccine the day that I did because she would have lost it or something, because they were so against it. They were really trying to enforce it.” (Interviewee D)*

Three participants attributed vaccine hesitancy to rumors of microchips being implanted through the vaccine to control people’s lives. One participant cited watching a YouTube video by a religious leader linking the vaccine to biblical references of 666 that correlated the COVID-19 pandemic and vaccine to, *“the belief of the devil taking over and how the transition would be”*. *(Interviewee D)*

Some participants pursued vaccination despite their communities’ prevailing negative beliefs against vaccination:

*“I trust my healthcare system so much even though my church community and family were against me getting the vaccine.” (Interviewee E)*

Conversely, one participant reported that their religious institution encouraged voluntary COVID-19 vaccination (Table 2). Their religious community actively combated vaccine hesitancy by holding information sessions in church and online and setting up a church-based vaccine clinic that provided information before walk-in immunizations to improve accessibility, especially for seniors (Table 2).

> *“There should be consultation on community, the leaders, because we are culturally different. But there are certain factors to be considered like the culture, … the way they live, the standard of living they have… the community leader knows this pretty well among his community. And maybe diversify the approach … I believe there should be a consultation among the different communities.” (Interviewee F)*

#### 2. Accessibility

Vaccine accessibility emerged as a major theme, with participants highlighting two key areas: access to reliable COVID-19 vaccine information and access to the vaccine itself.

### Access to COVID-19 Vaccine Information

Language barriers significantly affected access to credible vaccine information. Five participants cited language as a major barrier either for themselves or their community members (Table 3).

**Table 3.**
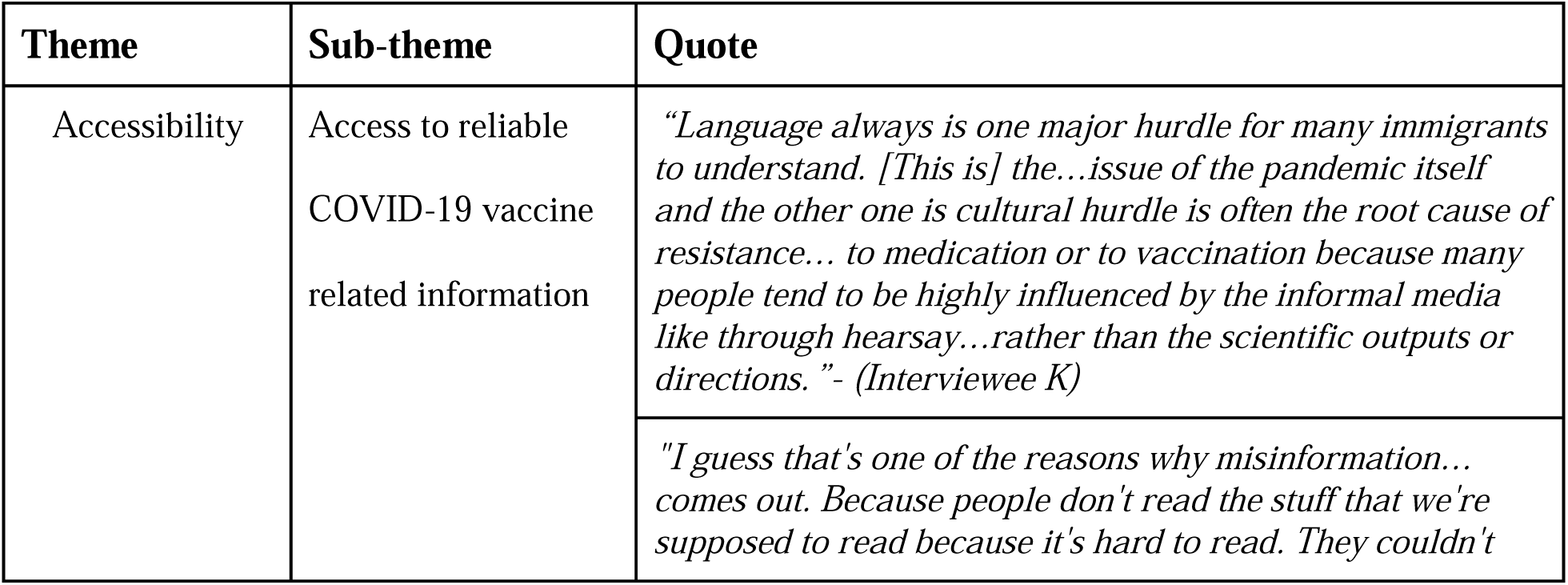
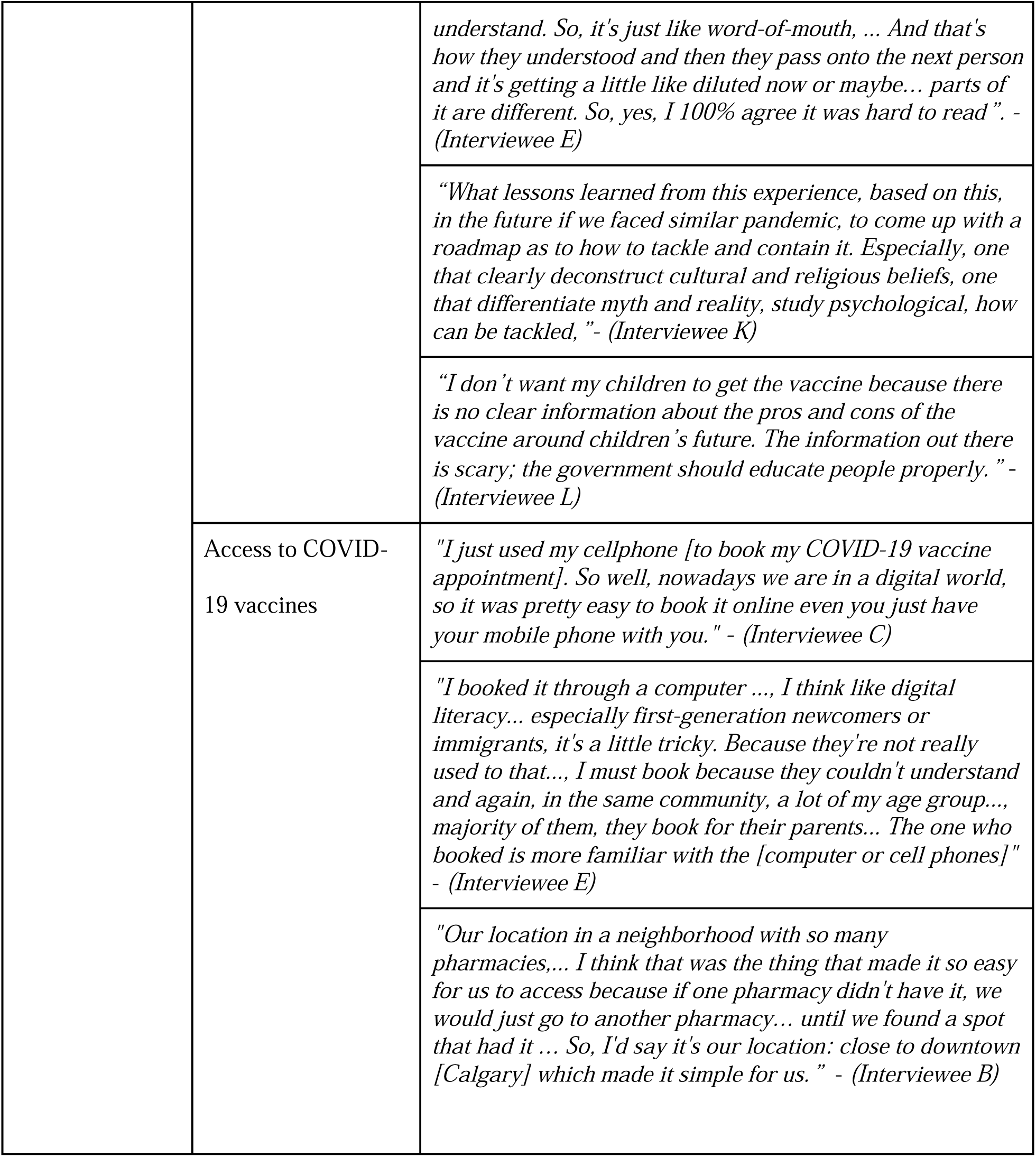
Access to credible COVID-19 vaccine related information and vaccines.

Participants mentioned that most available vaccine information was in high-level English, without readily available translations, making it difficult for vaccine-hesitant newcomers to understand health guidance. One participant mentioned that vaccine-hesitant family members did not read COVID-19 vaccine information provided by the health system because it was difficult to read. Another, who described themselves as English proficient stated that even after reading articles multiple times, they still could not understand them.

> *“Principally, the language barrier [played a role in vaccine hesitancy] that was my case because I have to translate everything that [the provincial health system] was putting on the website […]. I think the bigger problem is the language barrier … , seeing because well, people cannot understand and when they don’t understand, well they are not cooperating”. (Interviewee A)*

### Preferred formats for receiving COVID-19 vaccine information

Participants preferred credible, evidence-based vaccine information in first their language, using simple, non-technical terminology. Preferred formats included animated videos, roadmaps, webinars, and reports consolidated on websites or Facebook pages. They also preferred information was shared via schools, churches, community and newcomer organizations for credibility.

> *“In terms of accessibility, language matters. Especially with newcomers, plain, easy language.” (Interviewee E)*

This participant elaborated that constantly translating high-level information in English into simplified English, then into their first language was, “*exhausting.*”

### Access to COVID-19 Vaccines

Participants also described geographic and transportation barriers that limited vaccine access, as well as complex vaccine rollout stages implemented in Alberta. Some reported traveling long distances to find vaccination sites, particularly those living outside central urban areas (Table 3). For example, one participant stated that they had to drive to another city 150km away to receive the COVID-19 vaccine.

> *I know that if we didn’t live downtown and because we don’t have a car, I know that that would have definitely put us at a disadvantage because, like I said, other people were driving very long distances just to find a pharmacy where they could book a vaccine.” (Interviewee B)*

> *“During that time, the accessibility of vaccines was not inclusive. So even though I am convinced to get the vaccine, I have to drive 1 hour and 30 minutes to take the first dose.” (Interviewee G)*

Digital literacy and digital access played crucial roles in navigating the vaccine appointment system and identifying immunization clinics. Half of the study participants reported using their digital skills for themselves and family members to receive one or more vaccine doses. Some newcomers relied on English-proficient family members, while others received assistance from newcomer serving community organizations that advocated for and supported newcomer communities with booking and accessing vaccines.

> *“You need to book an appointment online, which demands basic computer skills; most people I know from our community don’t have that.” (Interviewee H)*

Community walk-in vaccine clinics were other important interventions for those facing vaccination barriers. Three participants mentioned using walk-in vaccine clinics to receive one or more doses (Table 3).

> *“The second one, I didn’t book an appointment, it was like walk-in. I was downtown and I found at - I think it was [NAME] building there was like, you don’t have to book an appointment, you can take your second dose… It was so organized so good. I didn’t have any issues.” (Interviewee I)*

Participants reported changes in vaccine access between first, second, and booster doses. Initially, the vaccine was available only to specific groups, with long wait times depending on eligibility. Although the vaccine eventually became available to all, some groups still faced access barriers, particularly for pediatric vaccinations (<12 years old). Four participants were surprised when comparing the lack of accessibility and availability of vaccines in their home countries to the ease they have experienced in Alberta.

> *“If we only talk about people who want the vaccine, they’re not sure. Living in a situation where it’s not accessible to you like in other countries. Here in Canada, it’s easy. If you really wanted it, you can get it at any pharmacy. If we talk to communities outside of Canada like in Africa, that’s where the problem is. They might not even [be able to] get it.” (Interviewee D)*

Despite this, participants felt that lack of clear information and poor accessibility caused them to delay or avoid vaccinating their children.

> *“The reason we decided not to vaccinate our son is because we didn’t know what was going on. My spouse and my son still didn’t get vaccinated and so far, nothing happened…and I hope nothing will happen later.” (Interviewee J)*

#### 3. Vaccine Acceptance

We identified two distinct groups based on their motivations for receiving COVID-19 vaccines: voluntary vaccination and coerced vaccination.

### Voluntary Vaccination

This group consisted of individuals motivated to voluntarily receive vaccinations to protect themselves and their communities. They understood the pandemic’s severity and believed vaccination would help reduce viral transmission. One participant described initially hesitating but ultimately trusting the vaccine after experiencing minimal reaction to their first two doses and trusted the vaccine manufacturing company (Table 4).

> “I had a good reaction and I trust the company who had made it. Like for me for instance, it was Pfizer so I 100% trust Pfizer … because they are one of the reputable companies when it comes to making medicines.” *(Interviewee C)*

**Table 4.** Vaccine acceptance: voluntary vs. coerced.

| Theme | Sub-Theme | Quote |
| --- | --- | --- |
| Vaccine Acceptance | Voluntary vaccination | *"My friends got COVID, my partner's family got COVID, he got COVID, so of course I was scared. ... That's part of the* |
|  |  | *reason why I didn't hesitate to get the vaccine. Because I – it's happening now in my community, my close circle community." - (Interviewee E)* |
|  |  | *"For my experience for the vaccine, honestly, I heard from the - when they were talking in the TV shows about the vaccine and how it's important and ... the meetings..., at the church..., talking about the vaccine and its importance and how we can take it to help each other to stop this pandemic. We have to work with each other, so I was so excited [to take the COVID-19 vaccine] but actually I was forced to take the vaccine." - (Interviewee I)* |
|  | Coerced vaccination | *"well personally, if I'm being honest with you, there's only really few people that got the vaccine in my community and most people that got the vaccine, in my community were younger than 20 - no, no, no - younger than 30, you know? So that's people that they're still chasing their dreams, chasing their careers and with the COVID-19 blocking everything, they had no choice because for some of us, we're at school and stuff and we still need it to keep up that lifestyle because we're still in the early age so there was almost no choice for us to get it." - (Interviewee D)* |
|  |  | *"He [family doctor] told me, it's my choice now because I'm suffering, I'm having complications [from the first dose of the COVID-19 vaccine]. So, he told me it's your choice. But, as I told you, I didn't have a choice because if I didn't get the second dose, I have to leave the job. And they [employer] scared us there, they told us, 'You know, everywhere you will go, it will be the same. You have to take the second dose at least to find a job'". - (Interviewee I)* |

**Table 5:**
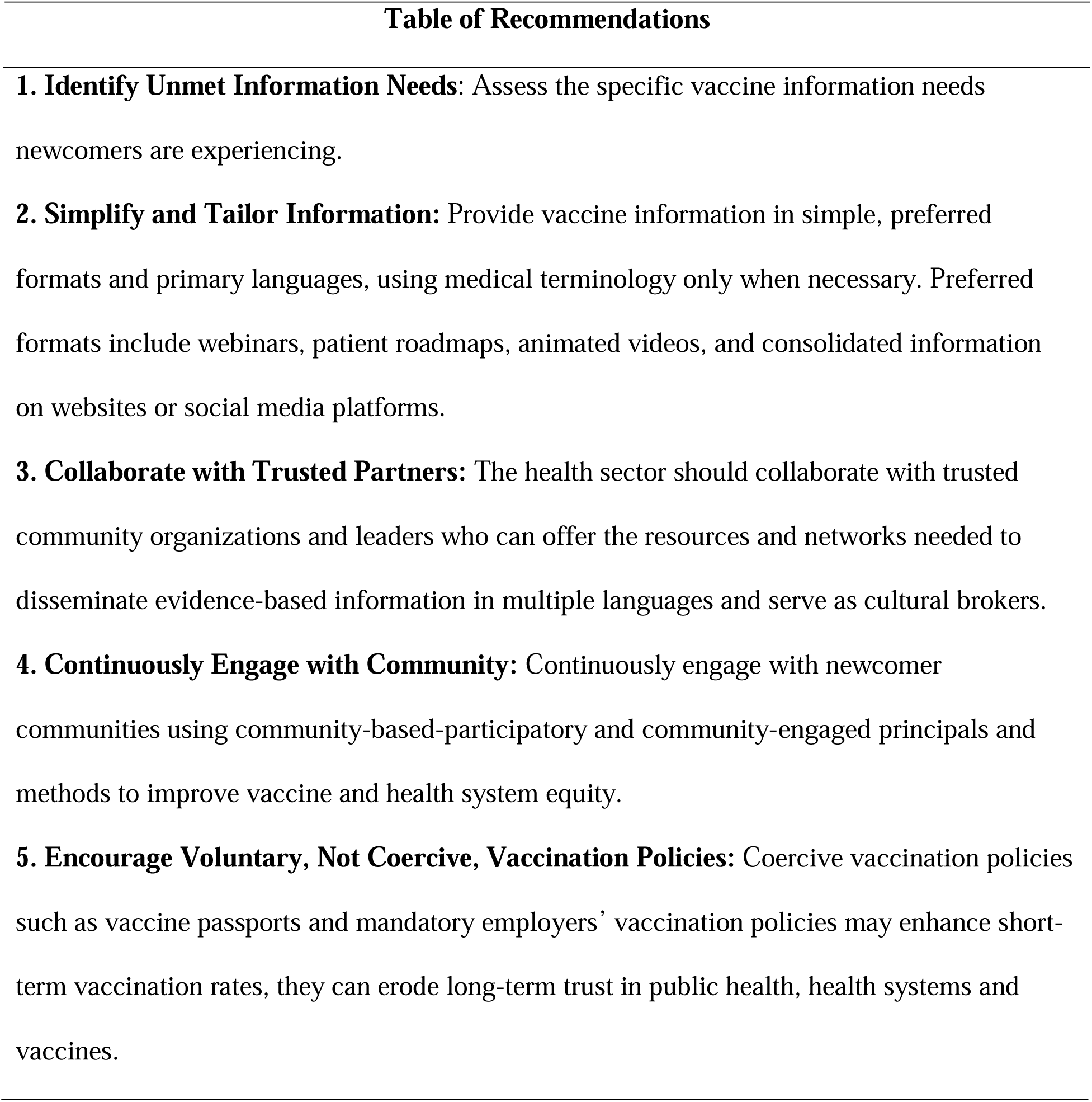
Key Recommendations to Improve Vaccine Acceptance and Reduce Vaccine Hesitancy Among Newcomer Communities.

### Coerced Vaccination

In contrast, three participants felt forced by their employers to receive the COVID-19 vaccine, fearing job loss during already uncertain times. They described feeling they had no choice but to get vaccinated to support their families, to pay rent, and to buy food. As COVID-19 restrictions lifted, some vaccinated individuals reported regretting their decisions due to community controversy and misleading information. Some felt overwhelmed when considering the second dose. Others reported anger and resentment, believing they were deceived.

> *“I still feel deceived. I feel like I was lied to if I’m being honest with you because I feel like the people who didn’t take [the] vaccine and stand on their feet, they won in a way. Now they can go anywhere with us and stuff like that. Things have slowed down. I feel like I have been deceived…” (Interviewee D)*

## Discussion

This community-based participatory research (CBPR) study investigated the experiences of newcomers in Alberta accessing COVID-19 vaccine information and vaccines during the final year before the World Health Organization declared the COVID-19 pandemic was no longer a global public health emergency (World Health Organization, n.d.). We identified three main themes, each with two sub-themes: vaccine hesitancy, vaccine accessibility, and vaccine acceptance. Our findings suggest newcomers’ vaccine hesitancy was influenced by cultural and religious beliefs, language barriers and mistrust due to misinformation. Participants faced difficulties discerning trustworthy information, especially when it was not available in their first language; thus, increasing hesitancy. Participants also felt stigmatized from their community members when they took vaccines, and they felt their community members questioned their reasoning of taking the vaccine. Similarly, vaccine accessibility was hindered by barriers such as language, digital literacy, geography and transportation. The study identified vaccine acceptance varied starkly between participants who voluntarily chose to be immunized, and those who felt coerced by employers. Importantly, perceived coerced vaccination emerged as a critical issue with potential long-term public health implications as this coercion led to greater mistrust in public health authorities and healthcare systems.

Our findings align with previous studies that highlight higher rates of vaccine hesitancy among newcomer communities when unable to access credible vaccine information through the healthcare system (Crawshaw et al., 2021). Similarly, our study participants reported experiencing fear and confusion when deciding on the safety of the COVID-19 vaccine, compounded by the global ‘infodemic’ of misinformation. The World Health Organization (WHO) declared a pandemic related ‘infodemic’ that created a global overabundance of pandemic-related mis and disinformation (World Health Organization, n.d.), leading to mistrust in health authorities that undermined the public health response, including vaccination against Sars-CoV2.

A prior study conducted across six Canadian provinces (Alberta, Ontario, Nova Scotia, British Columbia, Manitoba, and Saskatchewan) among Black communities found that misinformation increased fear and anxiety, leading to social divisions and stigma surrounding vaccine decisions (Kemei et al., 2023). In our study, participants also expressed concerns about losing community cohesion and the stigma associated with either accepting or declining the vaccine, which heightened anxiety and prevented individuals from receiving and promoting the vaccine. Like previous research, we found that language barriers and lack of reliable information in primary languages and preferred communication formats were recurring issues for newcomers, as reliable health information was often perceived as available only in advanced English (Smith et al., 2021). These language barriers intensified challenges, as participants had to translate information themselves, leading to further confusion. For example, our study participants described feeling overwhelmed by the amount of information circulating on news, social media, and government websites. To address this, study participants suggested that health systems should produce simplified health information in English as a Second Language level 5 for newcomers and leveraging preferred formats such as social media platforms, short videos and images.

The WHO states that healthcare should be available, accessible, acceptable, and of good quality as a human right (World Health Organization, n.d.). Language barriers prevent these goals from being fully met. A recent review of 84 studies by *Arya et al* concluded that for resettled refugees, employing professional interpreters can mitigate some of these challenges by improving communication, patient satisfaction, and ultimately, clinical outcomes (Arya et al., 2024). Our findings emphasize the importance of enhancing multilingual crisis communication to ensure newcomers and those with limited English proficiency can access reliable information for informed decision-making during a public health emergency (Arya et al., 2024).

During the COVID-19 pandemic, many healthcare systems initially overlooked the role of ethnocultural and religious leaders as trusted information sources and knowledge brokers (Campbell-Scherer et al., 2021). Including community members with lived experience as cultural brokers can help bridge the gap between healthcare systems and ethnocultural communities, improving vaccine communication and acceptance (Campbell-Scherer et al., 2021). Recognizing cultural beliefs and values is essential to achieve health equity and inclusion of diverse communities in health systems, especially in multicultural societies such as Canada. Potential solutions include collaborating with community leaders or international medical graduates with shared linguistic and cultural backgrounds to improve access to reliable vaccine–related information and reduce hesitancy (Smith et al., 2021). Religion also significantly influences vaccine decision-making in some newcomer communities. For example, previous studies have shown that faith–leaders have successfully increased vaccine confidence among ethnocultural minorities, especially those who mistrust government institutions (Song et al., 2024). One quarter of participants attributed vaccine hesitancy to religious beliefs. Collaborating with public health professionals to host information sessions and outreach vaccine clinics in faith-based settings can provide a trusted environment for accurate information and encourage vaccination (Song et al., 2024). These community-engaged approaches have proven effective in other settings, particularly in low- and middle-income countries, where faith communities activated volunteer networks to facilitate vaccine sign-ups and encourage attendance at vaccination appointments (Song et al., 2024).

### Accessibility to Vaccines

Various barriers faced by newcomer communities, including, digital literacy, language, geography and transportation, reduced vaccine access via different mechanisms. Digital literacy defined as “the ability to access, manage, understand, integrate, communicate, evaluate, and create information safely and appropriately through digital devices and networked technologies for participation in economic and social life” (Canadian Association of Research Libraries, n.d.) was critical for navigating online vaccine appointment systems. Complicated English-only online or telephonic vaccination booking protocols pose major barriers for newcomers with limited language proficiency (Arya et al., 2024). Aligned with previous studies, English proficient participants reported often acting as interpreters for family and other community members to help successfully navigate health systems (Katz, 2014). Participants with higher digital proficiency also found booking appointments easier; however, despite this, reported long wait times due to staged population eligibility during the pandemic as significant barriers. Geographic and transportation barriers, requiring newcomers to traverse long distances due to limited vaccine availability further limited vaccine access (Aylsworth et al., 2022). These barriers almost certainly exacerbate existing vaccine inequities among low-income newcomers or those living in rural settings (Holdbrook et al., 2023).

In keeping with other community-based participatory research (CBPR), our study participants identified multiple innovative solutions to help overcome existing vaccination barriers for newcomer communities. For example, participants highlighted low-barrier, community-based walk-in vaccine clinics as essential and preferred alternatives to tedious appointment-based clinics in distant locations (Holdbrook et al., 2023). Further, leveraging recent advances in artificial intelligence-enabled translation tools may provide other potential solutions to address language barriers. For example, the Canadian Refugee Healthcare System Atlas offers real-time multilingual navigation support for refugee health and settlement services (Refugee Health YYC, n.d.).

### Vaccine acceptance

Our findings regarding vaccine acceptance reinforce the existing literature, showing that a primary motivator for vaccination was the desire to protect oneself and one’s community members (Betsch et al., 2018). Those who were comfortable the vaccine would provide health benefits to themselves and their families, believed it was the right decision and were confident in their choice.

On the other hand, our study also highlights a group of newcomers who perceived they were coerced to receive vaccines, despite their concerns. These participants who felt compelled to vaccinate due to employer mandates, expressed uncertainty, and even regret, about their decision. They described fear of job loss as a primary motivator for being immunized. Although coerced vaccination has been associated with vaccine hesitancy in other jurisdictions (Deal et al., 2024), our participants described feeling unsafe when receiving the vaccine and wished they had more time to make informed decisions.

Vaccine acceptance among newcomers is more complex than anticipated, and deeper conversations with newcomers who felt coerced are necessary to fully understand their experiences. Understanding this complexity remains a particularly urgent global public health concern, given the worldwide decline in routine vaccinations and vaccine acceptance since the COVID-19 pandemic (UNICEF Canada, 2023). In Canada, this has led to re–emergence of vaccine preventable disease outbreaks such as measles, and polio, with large unvaccinated populations potentially at-risk (UNICEF Canada, 2023). These trends in reduced post-pandemic routine vaccinations, further highlight the potential public health risks and importance of applying the key pandemic learnings today (UNICEF Canada, 2023).

### Limitations

Our study has limitations. First, the study involved only three participants who had no prior relationship with the Community Scholars team. The CSs and research team were involved in COVID-19 vaccine outreach and public advocacy throughout the pandemic which may have biased participant’s study recruitment. However, immunized participants also shared their community’s challenges and insights about vaccine hesitancy based on their daily encounters and conversations. Future research could benefit from involving neutral third parties to improve recruitment strategies and reach a broader population. Second, the short timeline between ethics approval and PaCER course completion limited the ability to capture a wider scope and diversity of participants. Future research should focus on targeting communities more representative of newcomer communities in Alberta and purposively sampling underrepresented communities. Third, while the study recruited twelve participants representing different ethnocultural communities and CSs facilitated data collection in six primary languages, real-time translation for other commonly spoken primary languages was not offered. Future work should consider expanding the CS team to include other languages and newcomer groups. Expanding the research team’s participation in data collection would also facilitate better communication and reduce translation burdens.

### Conclusion

This study highlights the unique barriers newcomer communities in Alberta faced during the COVID-19 vaccine rollout, including language barriers, competing cultural and religious beliefs, misinformation, and access issues. These barriers were often overlooked, leaving many newcomers struggling to access and accept vaccines. The PaCER approach effectively engaged these communities, amplifying their voices and identifying gaps in public health outreach. Our findings show that healthcare systems must prioritize inclusive, culturally responsive strategies to better serve diverse newcomer populations. As routine vaccination rates fall and immigration levels reach record highs in post-pandemic Canada, public health programs must adapt to improve accessibility, provide information in multiple languages, and engage trusted newcomer community and faith leaders. These actions are critical for rebuilding trust and increasing vaccine uptake.

Finally, this research underscores the importance of making healthcare services inclusive and welcoming for all. By actively involving newcomers in designing vaccination programs, public health officials can reduce barriers, prevent outbreaks of vaccine-preventable diseases, and build a more resilient healthcare system. Empowering newcomers to view healthcare as safe and inclusive is vital to foster long-term trust and community well-being.

## Supporting information

COREQ Checklist

GRIPP Checklist

## Data Availability

Study data are available upon request.

## Declarations

### Ethics approval

This study was approved by the University of Calgary, Conjoint Health Research Ethics Board (CHREB) REB21-2060.

### Consent to participate

All study participants provided written or recorded oral consent

### Consent for publication

All study participants provided written or recorded oral consent to publish study findings

### Availability of data and material

Study data are available upon request.

### Code availability

Not applicable

### Author Contributions

MA, ERC, MY, AS, MY, IN, GEF, MS, DS, KP and LH led the study concept and design. MA, ERC, MY, AS, and MY actively supported data collection, data analysis, and manuscript preparation. MA, ERC, MY, AS, MY, IN, and LH provided support with writing. MA, ERC, MY, LH, IN, MS, DS, GEF, and MYE supported revising, editing, and subject-matter expertise. GEF takes responsibility for the final manuscript and provided scientific oversight through. All authors approved the final manuscript.

## Acknowledgements

We would like to thank the study participants for their vulnerability and enthusiasm for joining this study. We would also like to express our immense gratitude to Ingrid Nielssen and Dr. Maria Santana for their support in guiding us through our project and for providing opportunities to showcase our work. We would also like to thank the entire University of Calgary Refugee Health YYC Research team for their collective support and encouragement throughout the study process.

## Contributions to Knowledge

### What does this study add to existing knowledge?

- This study highlights potentially widespread unintended negative consequences of the pandemic vaccination campaign among newcomers, including COVID-19 vaccine regret and loss of trust in public health and healthcare officials.
- Our study adds to the post-pandemic assessment of broadly implemented vaccine mandate policies, and how they may help increase uptake of vaccines in short term while simultaneously decreasing trust in public health and health systems and government if they are poorly communicated to newcomer communities or pertinent information is not accessible to newcomers.
- Our study reinforces the importance of community based participatory research during health crises to better understand and overcome institutional mistrust and to create comprehensive public health outreach strategy for newcomer communities.

### What are the key implications for public health interventions, practice, or policy?

- Utilizing community-based participatory and patient-oriented research methods enables individuals with lived experience to co-design realistic solutions and policies within their communities. These methods are low-cost and transferable to other jurisdictions across culturally diverse communities in Canada.
- Our findings provide context-specific and universal recommendations for vaccination outreach strategies, that address vaccine hesitancy among newcomer communities, and can help newcomers navigate provincial healthcare more equitably.
- Better understanding how newcomer communities view, interpret, and want to engage with public health vaccination campaigns can inform engagement strategies for routine vaccination and other public health priorities such as cancer screening and health promotion.

