## Supplementary Materials for "Vaccine Hesitancy, Coercion and Regret: Post-Pandemic Lessons on COVID-19 Vaccine Policies and Outreach among Newcomer Communities in Alberta, Canada"

### **eFigure 1: Recruitment Poster**

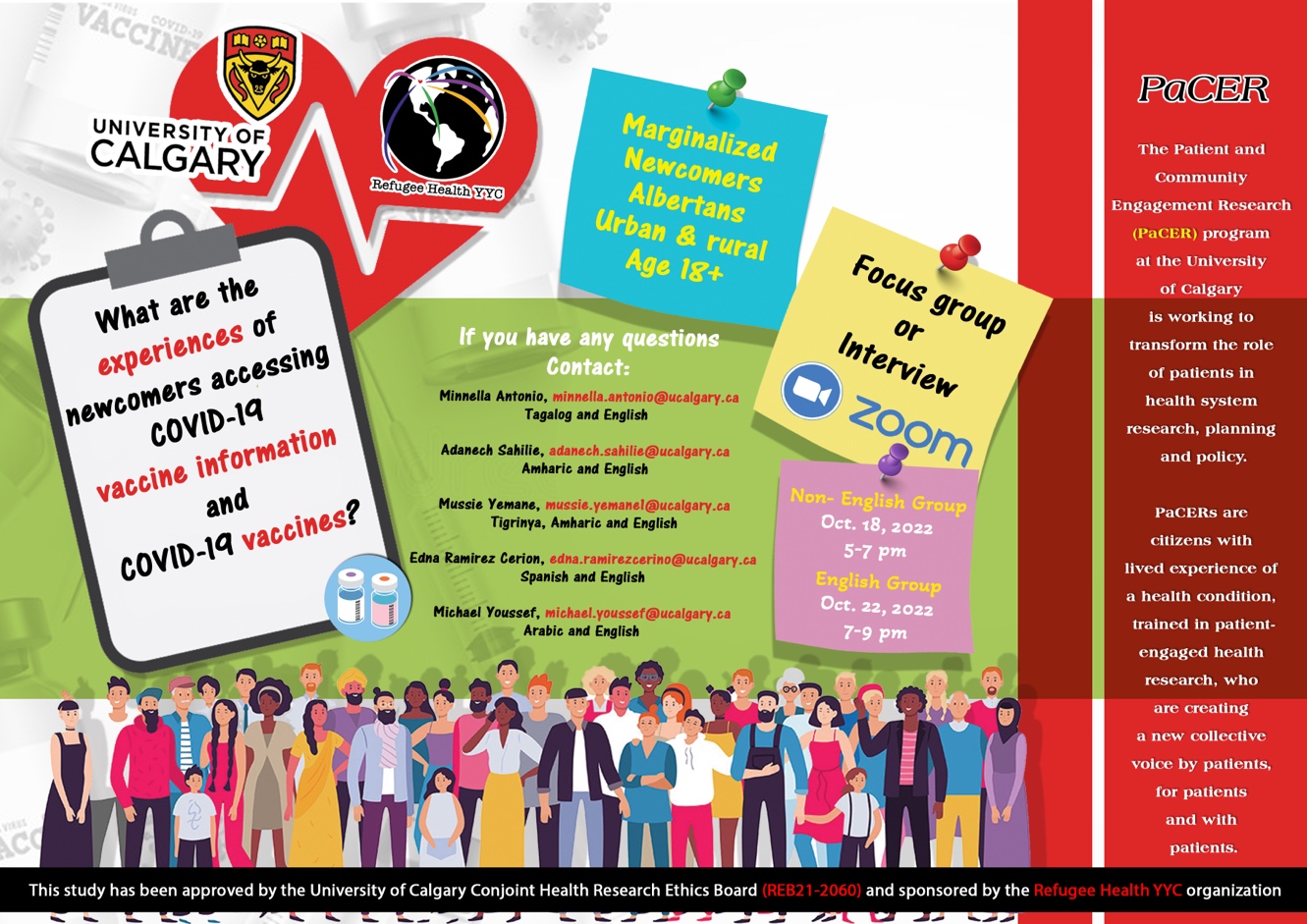

### **eTable 1: Question guide for COLLECT and REFLECT focus groups**

| **COLLECT focus group** | **COLLECT interview** |
| --- | --- |
| What is your name, age, gender, ethnicity, where you’re from and vaccine status? | Please introduce yourself |
| What is your favorite food? | How did you hear about our study? |
| How did you hear about the study? | How do you perceive the health of your community/your health after getting the vaccine?  Prompts to include: What was the impact of COVID-19 vaccine in your community/personal life/family? Did you have the opportunity to decide if you wanted to get vaccinated? How were you feeling with having the option/ not having the option to get the vaccine? |
| How do you perceive the health of your community/your health after COVID-19 pandemic started?  Prompts to include: What was the impact of COVID-19 in your community? What was the impact of COVID-19 in your personal life/family? | If you got the COVID-19 vaccine, where did you get it and what was your experience?  Prompts to include: Where did you get the vaccine? When? If you have not gotten the vaccine, would you like to share your thought about it OR reasons not to get it? |
| If you got the COVID-19 vaccine, where did you get it and what was your experience?  Prompts to include: Where did you get the vaccine? When? If you have not gotten the vaccine, would you like to share your thought about it OR reasons not to get it? | Where do you get information to COVID-19 vaccine?  Prompts include: Did you trust that information? Why or why not? Examples include Newspaper, social media (ex. Facebook, TikTok, Instagram, YouTube, WhatsApp), Television, Radio, Religious Institute, Word of Mouth (friends or family), Phone Calls, Podcast, Video, others. |
| Where do you get information about the COVID-19 vaccine?  Prompts include: Did you trust that information? Why or why not? Examples include Newspaper, social media (ex. Facebook, TikTok, Instagram, YouTube, WhatsApp), Television, Radio, Religious Institute, Word of Mouth (friends or family), Phone Calls, Podcast, Video, others. | How accessible to you feel the COVID-19 vaccine was to your community / age group?  Prompts to include: Could you please talk about what made it easier to get the vaccine OR what made it harder to get the vaccine? What was your experience while aiming to access and understand the information provided before getting the vaccine? |
| How accessible do you feel the COVID-19 vaccine was to your community / age group?  Prompts to include: Could you please talk about what made it easier to get the vaccine OR what made it harder to get the vaccine? What was your experience while aiming to access and understand the information provided before getting the vaccine? | Do you think some communities were able to access the COVID-19 vaccines and other were not?  Prompts to include: What has your experience been? |
| Do you think some communities were able to access the COVID-19 vaccines and others were not?  Prompts to include: What has your experience been? | Do you have any recommendations on how to access the vaccine? |
| Do you have any recommendations on how to access the vaccine? | What kind of outcomes do you expect to see from our research? |
| What kind of outcomes do you expect to see from our research? |  |

### **eTable 2: Top 20 codes used in data analysis**

| **Codes** | **Sum of Count of Codes** |
| --- | --- |
| Trusted Sources Of Information | 69 |
| Vaccine Accessibility | 54 |
| Vaccine Experience | 33 |
| Compare To Home Country | 28 |
| Easy | 28 |
| Trust In Healthcare System | 24 |
| Availability | 24 |
| Vaccine Hesitancy | 22 |
| Language Barrier | 22 |
| Workplace Experience | 18 |
| Mental Health | 17 |
| Sources Of Information | 16 |
| Mandatory Vaccination | 16 |
| Lack Of Trust In The Vaccine | 16 |
| Protect Community | 16 |
| Vaccine Acceptance | 15 |
| Shunned For Getting The Vaccine | 14 |
| Final Recommendation | 14 |
| Fear | 14 |
| Covid Experience | 14 |

### **eTable 3: Question Guide for REFLECT focus group**

**REFLECT**

Do you have any recommendations on how to access the vaccine?

Were themes captured correctly?

What have we missed?

How would you prefer to learn about the outcome of this research?

What language would you like us to share our report in?

What sources of information do you trust the most and why? Is there any way you can tell whether the source of information is reliable or not?

Do you think language barrier, cultural, socio-economic status and religious background played a role for vaccine hesitancy?

If a new pandemic arises, how do you think AHS or the government should handle it? What are their recommendations not to fall in the same trap again?

Any suggestions you have to make the vaccine more accessible?

How can accessibility be improved in the future?

What worked, what didn’t work and what would you do differently?

**eTable4:** COREQ (Consolidated criteria for Reporting Qualitative research) Checklist

**eTable5:** GRIPP (Guidance for Reporting Involvement of Patients and the Public) Checklist
