## Supplementary material for "Vaccine Hesitancy, Coercion and Regret: Post-Pandemic Lessons on COVID-19 Vaccine Policies and Outreach among Newcomer Communities in Alberta, Canada": GRIPP Checklist

**eTable 5: Guidance for Reporting Involvement of Patients and the Public (GRIPP)**

| Section and Topic | Item | Reported on Page No |
| --- | --- | --- |
| 1. Aims | \| Report the aim of PPI in the study | 5 |
| 2. Methods | Provide a clear description of methods used for PPI in the study \| | 6-8 |
| 3. Study Results | Outcomes: Report the results of PPI in the study, including both positive and negative outcomes | 10-17 |
| 4. Discussion and conclusions | Outcomes: Comment on the extent to which PPI influenced the study overall. Describe positive and negative effects | 18-23 |
| 5. Critical perspective | Comment critically on the PPI in the study, reflecting on the things that went well and those that did not, so others can learn from this experience | 23 |
